# Physiotherapy service provision in a specialist adult cystic fibrosis service: a pre-post design study with the inclusion of an allied health assistant

**DOI:** 10.1101/2020.11.16.20232256

**Authors:** Kathleen Hall, Lyndal Maxwell, Robyn Cobb, Michael Steele, Rebecca Chambers, Mark Roll, Scott C Bell, Suzanne Kuys

**Affiliations:** School of Allied Health, Faculty of Health Sciences, Australian Catholic University, 1100 Nudgee Road, Banyo, QLD, 4014, Australia; Physiotherapy, The Prince Charles Hospital, 627 Rode Road, Chermside, QLD 4032, Australia; Adult Cystic Fibrosis Centre, The Prince Charles Hospital, 627 Rode Road, Chermside, QLD 4032, Australia; Translational Research Institute, 37 Kent Street, Woolloongabba, QLD, 4102, Australia; Children’s Health Research Centre, The University of Queensland, Herston QLD 4006, Australia; Nursing Research and Practice Development Centre, The Prince Charles Hospital, 627 Rode Road, Chermside, QLD 4032, Australia

**Keywords:** Allied Health Assistants, Cystic Fibrosis, Physiotherapy, Health Workforce, Scope of Practice, Delivery of Healthcare

## Abstract

**Question(s):** What is the impact of including an allied health assistant (AHA) role on physiotherapy service delivery in terms of service provision, scope of practice and skill mix changes in an acute respiratory service?

**Design:** A pragmatic pre-post design study examined physiotherapy services across two three-month periods: current service delivery [P1] and current service delivery plus AHA [P2].

**Outcome measures:** Clinical and non-clinical activity contributing to physiotherapy services delivery quantified as number, type and duration (per day) of all staff activity, and categorised for skill level (AHA, junior, senior).

**Results:** Overall physiotherapy service delivery increased in P2 compared to P1 (n=4730 vs n=3048). Physiotherapists undertook fewer respiratory (p < 0.001) and exercise treatments (p < 0.001) but increased patient reviews for inpatients (p < 0.001) and at multidisciplinary clinics in P2 (56% vs 76%, p < 0.01). The AHA accounted for 20% of all service provision. AHA activity comprised mainly non-direct clinical care including oversight of respiratory equipment use (e.g. supply, set-up, cleaning, loan audits) and other patient related administrative tasks associated with delegation handovers, supervision and clinical documentation (72%) and delegated supervision of established respiratory (5%) and exercise treatments (10%) and delegated exercise tests (3%). The AHA completed most of the exercise tests (n = 25). AHA non-direct clinical tasks included departmental management activities such as statistics and ongoing training (11%). No adverse events were reported.

**Conclusion:** Inclusion of an AHA in an acute respiratory care service changed physiotherapy service provision. The AHA completed delegated routine clinical and non-clinical tasks. Physiotherapists increased clinic activity and annual reviews. Including an AHA role offers safe and sustainable options for enhancing physiotherapy service provision in acute respiratory care services.

## INTRODUCTION

People with cystic fibrosis (CF) are living longer, thereby increasing both patient numbers and complexity of care.^1,2^ Demand for services is expected to further increase, with forecasts suggesting the number of adults living with CF increasing by 75% between 2015 and 2025^3,4^ Notably these estimates don’t incorporate predicted increases in survival associated with the addition of cystic fibrosis transmembrane conductance regulator modulator therapies.^1^ Multidisciplinary and expert care is recommended by international guidelines.^5,6^ Previously we demonstrated the effect of limited physiotherapy staffing numbers and skill mix in a large tertiary CF centre, where current demand exceeded supply.^7^ Key aspects of CF physiotherapy treatment (respiratory and exercise treatments) met recommended guidelines.^7,8^ However, aspects of care such as exercise testing and detailed clinical care review did not.^7,8^ Many other adult CF services appear equally under resourced to deliver adult CF care^4^ and likely facing challenges to provide ongoing physiotherapy service delivery and future sustainability of care^1,4^.

Innovative strategies to manage increased service demand such as remodelling care delivery using allied health assistants (AHAs) has been recommended in areas of acute care but not for people with CF.^9-11^ AHAs have been identified as a potential cost-effective resource for health care delivery yet they appear to be underutilised.^12^ AHAs are well received by patients and can perform both clinical and non-clinical tasks,^13,14^ thereby improving efficiency and allowing allied health professionals (AHPs) to spend more time performing clinical care or other duties.^15,16^ Comprehensive information about establishing AHA roles and changes to physiotherapy practice associated with such roles in an acute respiratory clinical setting has not been reported.

We aimed to determine the impact of the inclusion of an AHA role on physiotherapy service delivery in an acute respiratory care setting (adult CF centre), in terms of service provision, scope of practice and skill mix changes.

## MATERIALS AND METHOD

### Design

A pragmatic pre-post design study was conducted at an adult CF centre to examine the delivery of physiotherapy services across two three-month periods: phase one (P1) [September – November 2015] and phase two (P2) [April – June 2016]. Data collection periods were chosen to avoid peak holiday (December to January) and clinical demand (July to August) to minimise variations in patient demand and complexity between phases. Physiotherapy staffing in both phases comprised two full time equivalent permanent senior CF physiotherapists and two full time equivalent junior rotational physiotherapists. A full time equivalent AHA role was included in phase two staffing. A workforce redesign tool, the Calderdale Framework, was used for the development of the AHA role.^17,18^ This framewrok engages staff in a staged sytematic approach to reviewing skill mix and developing new roles.^17,18^

All CF physiotherapy staff and the AHA were trained in the principles and practices of delegation.^19^ Physiotherapy staff delegated activities to an AHA who had been specifically trained in the knowledge and skills to undertake the activities safely.^19,20^ Training covered direct and non-direct clinical tasks (Supplement 1 Table A). Clinical tasks included six-minute walk tests, and supervision of established inhalation therapy, airway clearance and routine exercise treatments for stable inpatients as delegated by the physiotherapy staff. Non-direct clinical tasks included oversight of respiratory equipment use (including supply, cleaning, audit of loans) and general administrative duties.

### Data collection

Physiotherapy services for adults with CF, both inpatient and outpatient, were quantified during weekdays (usual business hours). Staff recorded all direct and non-direct clinical and non-clinical activity daily to quantify physiotherapy service delivery using a portable scanning system (Chappell Dean Pty Limited). Data included date, time, location (ward, multidisciplinary outpatient clinics), activity type using a predetermined code list,^7^ number and duration of each activity, and staffing level (junior, senior, AHA in phase 2).

Clinical data were collected for all patients with CF admitted to the hospital across both phases. The number of admissions and number of people with CF attending multidisciplinary outpatient clinics were recorded over each phase. Number and details of any documented clinical incidents or adverse events during any physiotherapy or AHA intervention were recorded using the hospital incident recording system^21^ for each phase. Staff and patient perceptions of the physiotherapy service during both phases were sought from all members of the specialist CF multidisciplinary team and all patients receiving physiotherapy care during both phases. Purpose designed surveys were developed and pilot tested with both participant groups. Patient surveys comprised 26 questions and staff surveys nine questions (Supplement 2). The surveys sought perceptions of the quality, effectiveness and accessibility of the physiotherapy service. A five-point Likert scale was used with open-ended response options also provided for participants to provide additional information.

### Outcome measures

The primary outcome was all clinical and non-clinical activity that contributed to the delivery of physiotherapy services. Physiotherapy services were described under three categories: service provision, scope of practice and skill level. Service provision is quantified as the numbers of inpatients admissions and outpatient attendances at multidisciplinary clinics. Scope of practice activity was quantified as number, type and duration (per day) of all staff activity, further categorised for skill level (AHA, junior, senior).^7^ Adverse events were described in terms of type and number.

### Data analysis

Demographic, service provision, scope of practice and skill mix data were analysed descriptively. Fisher’s exact tests were used to determine differences in service delivery, staff numbers and types of activities across phases. Independent t-tests were conducted to compare the number of activities per day and duration of activity type on the days these activities occurred between phases for all staff and between junior and senior staff. Clinical and demographic information from surveys were analysed descriptively. Mann-Whitney U tests were conducted to compare survey responses between phases. Open-ended responses were collated. Significance was defined as a p value < 0.05. SPSS v25 (IBM Corp., NY, USA) for all analyses.

## RESULTS

### Service provision

In P1, there were 113 inpatient admissions and 385 patient attendances at multidisciplinary outpatient clinics (henceforth called clinics). In P2, there were 111 inpatient admissions and 352 patient attendances at clinics. All inpatients across both phases received direct clinical care by the physiotherapy service. Physiotherapists saw a higher proportion of attendees at clinics in P2 (268 (76%) vs 215 (56%), absolute risk difference 20% (95% confidence interval 13 to 27).

### Scope of practice

Physiotherapy service activity (n, %) for all staff across the phases is described in Table 1. Overall, the physiotherapy service undertook more activity in P2 (n = 4730) compared to P1 (n = 3048).

**Table 1.** Number (percent total activity) of clinical and non-clinical care activities by all staff (physiotherapists and AHAs) across phase one and two. Comparisons between phases for all staff (number (%).

| Activity | All Staff |  | Fishers exact test (p) |
| --- | --- | --- | --- |
|  | Phase 1<br>n (%) | Phase 2<br>n (%) |  |
| *Clinical care: Direct* |  |  |  |
| Respiratory Treatment | 1058 (35) | 830 (18) | <0.001 |
| Exercise Treatment | 338 (11) | 350 (7) | <0.001 |
| Exercise Test | 20 (1) | 40 (1) | 0.426 |
| Multidisciplinary team clinics | 215 (7) | 268 (6) | 0.01 |
| Reviews | 79 (3) | 342 (7) | <0.001 |
| Other Treatment<sup>##</sup> | 29 (1) | 20 (0) | <0.01 |
| *Total direct clinical care* | *1739 (57)* | *1850 (39)* |  |
| *Clinical care: Non-direct* |  |  |  |
| Patient related documentation, communication and management<sup>###</sup> | 749 (25) | 1796 (38) | <0.001 |
| Equipment management<sup>####</sup> | 102 (3) | 273 (6) | <0.001 |
| *Total non-direct clinical care* | *851 (28)* | *2069 (44)* |  |
| *Total clinical care* | *2590 (85)* | *3919 (83)* |  |
| *Non Clinical care* |  |  |  |
| Management | 326 (11) | 587 (12) | 0.037 |
| Teaching & training | 128 (4) | 187 (4) | 0.56 |
| Research | 4 (0) | 37 (1) | <0.001 |
| *Total non-clinical care* | *458 (15)* | *811 (17)* |  |
| **Total Activity** | **3048** | **4730** |  |

Reviews include physiotherapy annual review assessment and/or detailed reviews of specific management;
<sup>##</sup>Other includes routine musculoskeletal and incontinence management and other clinical care activity not covered in other categories; <sup>###</sup>Patient related documentation and communication includes documentation of clinical care related to patients and all other clinical documentation related to patient care administration and other patient related clinical activities (handovers, weekly patient review meetings) not covered in other categories; <sup>####</sup>Equipment management includes time taken to manage (supply / setup / clean / order) patients respiratory / oxygen therapy equipment. Please refer to Hall K et al., 2020<sup>7</sup> for a full description of activity code inclusions.

### Physiotherapist activity

Table 2 presents the activity undertaken by physiotherapists and AHA for both phases. Overall, the number and percentage of clinical care activities undertaken by physiotherapists across the two phases was similar (85 vs 81%) with some differences between the phases for specific activities (Table 2). In P2, physiotherapists undertook fewer respiratory and exercise treatments and patient reviews increased from 79 to 342. Patient related clinical administrative tasks such as documentation, handovers, attendance at ward rounds and discussions within the multidisciplinary team increased from 25% in P1 to 36% in P2. Activity associated with managing patients’ equipment needs by the physiotherapists reduced in P2. Non-clinical care activities of research and management increased from P1 to P2. Teaching and training remained unchanged (Table 2).

**Table 2.** Number (percent total activity) of clinical and non-clinical care activities undertaken by physiotherapists and AHA for each phase.

| Activity | Phase 1 | Phase 2 |  |
| --- | --- | --- | --- |
|  | Physiotherapists | Physiotherapists | AHA |
|  | n (%) | n (%) | n (%) |
| *Clinical care: Direct* |  |  |  |
| Respiratory Treatment | 1058 (35) | 778 (21) *** | 52 (5) |
| Exercise Treatment | 338 (11) | 257 (7) *** | 93 (10) |
| Exercise Test | 20 (1) | 15 (0) | 25 (3) |
| Multidisciplinary team clinics | 215 (7) | 268 (7) ** | 0 |
| Reviews | 79 (3) | 342 (9) *** | 0 |
| Other Treatment | 29 (1) | 19 (1) * | 1 (0) |
| *Total direct clinical care* | *1739 (57)* | *1679 (45)* | *171 (18)* |
| *Clinical care: Non-direct* |  |  |  |
| Patient related documentation, communication and management | 749 (25) | 1363 (36) *** | 433 (45) |
| Equipment management | 102 (3) | 19 (1) *** | 254 (26) |
| *Total non-direct clinical care* | *851 (28)* | *1382 (37)* | *687 (72)* |
| *Total clinical care* | *2590 (85)* | *3061 (81)* | *858 (90)* |
| *Non Clinical care* |  |  |  |
| Management | 326 (11) | 485 (13) * | 102 (11) |
| Teaching & training | 128 (4) | 187 (5) | 0 |
| Research | 4 (0) | 37 (1) *** | 0 |
| *Total non-clinical care* | *458 (15)* | *709 (19)* | *102 (11)* |
| **Total Activity** | **3048** | **3770** | **960** |

\* p<0.05; \*\* p=0.01; \*\*\* p<0.001, p values based on Fisher's exact t test of the difference between phases

### AHA activity

The AHA completed 960 activities in P2, representing 20% of all physiotherapy service provision (Table 2). The majority of this was non-direct clinical care (n = 687, 72%), however delegated direct clinical activity including respiratory (n = 52 (5%) and exercise treatments (n = 93, 10%) occurred, contributing to the overall increase in numbers of exercise treatments undertaken by all staff in P2 (Table 2). The AHA completed 25 (3%) delegated exercise tests (Table 2).

### Time taken per activity by staff

The mean duration of each episode of activity per day for P1 and P2 for all staff is described in Table 3. Time spent on respiratory treatments increased by four minutes per episode in P2. Reduced time was spent on documentation, management and communication activities per episode in P2 (Table 3). Time spent on remaining activity episodes didn’t change (p > 0.05).

**Table 3:** Duration in minutes (mean (SD)) of each episode of activity per day of clinical and non-clinical care activities by all staff for each phase. Mean difference (95% confidence interval (CI)) between the two phases.

| Activity | Duration of each episode of activity per day (mins) |  |  |
| --- | --- | --- | --- |
|  | Phase 1<br>Mean (SD) | Phase 2<br>Mean (SD) | Mean difference<br>(95% CI)<br>P2 minus P1 |
| *Clinical care: Direct* |  |  |  |
| Respiratory Treatment | 34 (8) | 38 (4) | 4 (2 to 6) |
| Exercise Treatment | 41 (7) | 39 (6) | - 2 (-4 to 0) |
| Exercise Test | 32 (9) | 35 (7) | 3 (-2 to 8) |
| Multidisciplinary team clinics | 51 (29) | 53 (31) | 2 (-11 to 14) |
| Reviews | 42 (10) | 41 (11) | -1 (-5 to 4) |
| Other Treatment | 24 (6) | 32 (13) | 8 (1 to 16) |
| *Clinical care: Non-direct* |  |  |  |
| Patient related documentation, communication and management | 25 (14) | 10 (5) | -15 (-18 to -11) |
| Equipment management | 27 (12) | 36 (15) | 9 (3 to 14) |
| *Non Clinical care* |  |  |  |
| Management | 50 (37) | 48 (12) | -2 (-12 to 8) |
| Teaching & training | 49 (17) | 40 (12) | -9 (-15 to -3) |
| Research | 65 (17) | 69 (65) | 4 (-62 to 71) |

### Skill mix

Overall junior physiotherapists undertook similar number (Table 4) and duration (Supplement 1 Table B) of direct clinical care activities in both phases. Direct clinical care activity increased for the number of reviews, and junior physiotherapists commenced non-clinical teaching and training activity in P2. No research activity for junior physiotherapists occurred in either phase (Table 4).

**Table 4.** Number (mean (SD)) of clinical and non-clinical care activities per day undertaken by junior and senior physiotherapists across each phase.

| Activity | Junior physiotherapists |  |  | Senior physiotherapists |  |  |
| --- | --- | --- | --- | --- | --- | --- |
|  | Phase 1<br>Mean (SD) | Phase 2<br>Mean (SD) | Mean difference<br>(95% CI)<br>P2 minus P1 | Phase 1<br>Mean (SD) | Phase 2<br>Mean (SD) | Mean difference<br>(95% CI)<br>P2 minus P1 |
| *Clinical care: Direct* |  |  |  |  |  |  |
| Respiratory treatment | 8.3 (4.5) | 9.5 (3.3) | 1.3 (-0.1 to 2.7) | 8.3 (8.5) | 3.0 (2.8) | -5.3 (-7.5 to -3.0) |
| Exercise treatment | 4.4 (2.1) | 3.6 (2.0) | -0.8 (-1.5 to -0.1) | 0.9 (1.2) | 0.6 (0.7) | -0.3 (-0.7 to 0.0) |
| Exercise testing | 0.2 (0.5) | 0.2 (0.4) | 0.0 (-0.2 to -0.1) | 0.1 (0.4) | 0.1 (0.2) | 0.0 (-0.2 to 0.1) |
| Multidisciplinary team clinics | 0.2 (0.9) | 0.0 (0.1) | -0.2 (0.5 to -0.0) | 3.1 (3.2) | 4.3 (4.3) | 1.2 (-0.2 to 2.5) |
| Reviews | 0.2 (0.5) | 1.9 (1.7) | 1.6 (1.2 to 2.1) | 1.0 (1.3) | 3.7 (2.9) | 2.8 (1.9 to 3.6) |
| Other treatment | 0.4 (1.1) | 0.3 (0.5) | -0.1 (-0.4 to -0.2) | 0.1 (0.3) | 0.0 (0.2) | 0.0 (-0.1 to -0.1) |
| *Clinical care: Non-direct* |  |  |  |  |  |  |
| Patient documentation /communication/<br>management | 3.0 (3.7) | 8.0 (4.5) | 5.0 (3.8 to 6.2) | 2.9 (5.5) | 14.4 (10.1) | 11.5 (8.7 to 14.3) |
| Equipment management | 0.3 (0.6) | 0.2 (0.5) | -0.2 (-0.4 to -0.0) | 1.3 (2.7) | 0.1 (0.4) | -1.1 (-1.8 to -0.4) |
| *Non clinical care* |  |  |  |  |  |  |
| Management | 2.5 (2.0) | 4.7 (1.7) | 2.2 (1.5 to 2.8) | 2.6 (2.1) | 3.2 (1.7) | 0.6 (-0.1 to 1.2) |
| Teaching and training | 0* | 1.1 (1.5) | 1.1 (0.7 to 1.5) | 2.0 (1.5) | 1.9 (1.2) | -0.1 (-0.6 to -0.4) |
| Research | 0* | 0 | <sup>a</sup> | 0.1 (0.2) | 0.6 (0.6) | 0.5 (0.4 to 0.7) |

<sup>a</sup>. t cannot be computed because there were no data for at least one of the groups
\* Represents nil activity

Differences in most of the clinical care activities were observed for senior physiotherapists between the phases (Table 4). In P2, senior physiotherapists completed fewer respiratory and exercise treatments, however spent longer time per episode compared to P1 (Supplement 1 Table B). Senior physiotherapists completed the same number of exercise tests in P2 (Table 4), though approximately 16 minutes longer was spent completing each test (Supplement 1 Table B). Senior physiotherapists increased the number of inpatient reviews completed per day from 1.0 (SD1.3) in P1 to 3.7 (SD2.9) in P2 (Table 4). There was no difference in number or duration of non-clinical care activities for teaching and training and management for senior physiotherapists. Senior physiotherapists undertook more research activity in P2 (Table 4).

### Safety

No clinical incidents or adverse events associated with any physiotherapy or AHA patient intervention were reported to the investigators nor reported through the hospital clinical incidents system (PRIME)^21^ across P1 or P2.

### Perceptions of staff and patients

Eighteen (51%) and 17 (49%) staff responded to surveys during P1 and P2 respectively; 40% were allied health staff, 23% nursing and 29% medical staff. Sixty-three (35%) and 62 (36%) CF patients (53% male, 39% aged 36 years or older) receiving physiotherapy responded during P1 and P2 respectively. Staff (88%) were aware of the AHA working within the physiotherapy team during P2 and rated there was improved access to physiotherapy services for patients (p = 0.05) and greater ability of senior physiotherapy staff to engage in clinical care discussions and research (p < 0.05). Approximately two-thirds (62%) of patients reported the AHA had been involved in their care in P2, with 87% of respondents rating their physiotherapy care as good to excellent (Supplement 1 Figure A). In P1 76% agreed or strongly agreed their physiotherapy care was effective with different staff involved in their care, which increased to 88% of respondents in P2 (Figure 1). Overall, written responses were few with no negative comments associated with the AHA delivering provision of care. Respondents indicated that the care was ‘*still a high standard’* (participant X) and perceived the AHA as a ‘*good resource, interested and knowledgeable’* (participant Y).

**Figure 1:**
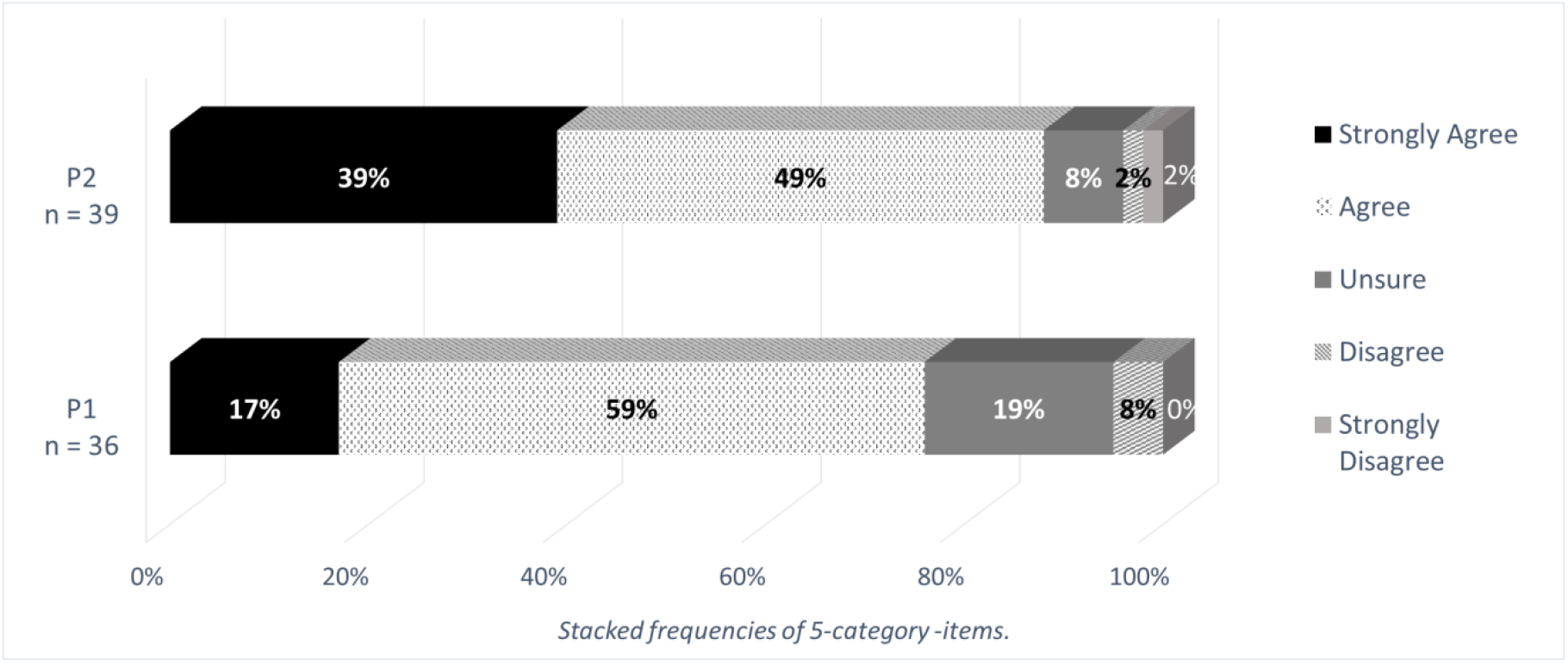
Perceived effectiveness of the physiotherapy care delivery in each phase of the study, rated on a 5-category Likert-type scale. Mean ranks: 43 for P1 and 33 for P2, Mann-Whitney U = 508, p = 0.024.

## DISCUSSION

This study describes the successful development and incorporation of an AHA role in an acute CF physiotherapy service. This redesign was an innovative approach to address service provision challenges^7^ associated with increasing age, numbers and complexity of care for adults with CF.^1^ Redesigning health service delivery, where change is directed towards skill mix reconfiguration and optimising healthcare team capabilities to increase workforce capacity and patient outcomes is well recognised.^9,11,12,22^

Overall access to physiotherapy services improved according to the multidisciplinary team. Additionally, more patients reported their care was effective in the second phase of the study. Approximately two-thirds of these patients reported the AHA had been involved in this care. Physiotherapists scope of practice changed, incorporating more advanced skills such as patient reviews and research. In conjunction with delegation of suitable tasks to the AHA, it would appear physiotherapy service delivery in this centre moved closer to benchmarking standards recommended in clinical practice guidelines, with increased exercise testing and physiotherapy activity in clinics.^7,8^ These findings describe a redesigned acute care respiratory physiotherapy service with increased capability, comprising a new skill mix of an AHA and junior and senior physiotherapists. Similar service delivery models have been shown to improve patients outcomes in previous studies.^13,14,22^

The AHA contributed 20% of overall physiotherapy service delivery with approximately 90% of their work providing direct and non-direct clinical care activity. Direct care included delegated respiratory and exercise treatments. In fact, most of the exercise tests in the second phase of the study were completed by the AHA. Previous reports of AHAs providing acute hospital ward-based physiotherapy care include delegated strengthening and balance exercises and mobilisation occurring on rehabilitation, orthopaedic and general medical wards^11,22^ and mobilisation of patients post abdominal surgery.^22^ This is the first time AHA workloads have been quantified for specific acute respiratory physiotherapy treatments to our knowledge. Of note, no patients reported their quality of care was compromised.

As a likely consequence of the new AHA role within the physiotherapy service, changes to physiotherapists’ scope of practice occurred. Some exercise treatments and the management of patients’ equipment appeared to have shifted to the AHA. Senior physiotherapists could complete more patient reviews, undertake more activity within the clinics and increase research activity. Junior physiotherapists undertook more advanced roles, including teaching and training, and some patient reviews. All physiotherapists increased their engagement in patient communication and management activity. Multidisciplinary team members felt the physiotherapy staff could contribute more to clinical care discussions. Clinical guidelines endorse the importance of physiotherapists being available for daily pre and post clinic meetings, inpatients discussion meetings, weekly multidisciplinary team case conferences and contributions to research and education meetings^1,5,8,28^ thus an AHA role is a possible strategy to optimise scope of practice for physiotherapists.

Barriers to the successful development and implementation of an AHA role within an acute care respiratory physiotherapy service were considered in the planning of this service redesign, in particular the ability to safely contribute to direct respiratory clinical care delivery. It is well documented that barriers to successful AHA role development include pre-existing perceptions of both physiotherapists and AHAs about these roles.^14-16^ Other barriers about the AHA role include lack of clarity regarding the specific tasks to be performed, the need for preparation and training, and an understanding by all staff about the level of accountability and responsibility for treatments undertaken by the AHA, which requires training for all staff in supervision and delegation practices.^11,14,15,22^ To address these issues the Calderdale Framework^18^ was used to develop the AHA role, with a focus on supporting skill mix redesign and mitigating potential risk.

This workforce redesign tool was specifically chosen as has been successfully used in the implementation of AHA roles,^26^ is patient-focused and engages both the AHAs and the physiotherapists in the seven-step facilitated process.^18^ Using the Calderdale Framework analysis tools and a trained facilitator^17,19^ AHAs and physiotherapists worked together to determine clinical and non-clinical tasks to be included in the new AHA role. AHA training was comprehensive and followed a taught, modelled, competent methodology.^27^ Competency assessment, clinical governance processes, and a procedure for documentation and feedback to delegating physiotherapists after task completion were developed. All staff completed a structured delegation training process.^19^ This training outlined the level of accountability and responsibility for both the physiotherapists and AHAs when supervising and handing over clinical tasks to AHAs.^15^

It is likely therefore, that the positive outcomes reported in this study can be attributed to the use of a comprehensive workforce development tool to develop and implement the AHA role. All staff appeared to be engaged in activity at appropriate scope, which included the development of an appropriate scope of practice for the AHA and then physiotherapy staff undertaking more advanced scope of practice activities required to deliver care to this complex patient group.^11,18^

Another aim of the using the workforce redesign tool was to mitigate potential risk. This appears to have been achieved. The delegated clinical treatments undertaken by the AHA in this study appear to be safe, as no major adverse clinical events were reported in the hospital’s clinical incident documentation system. Previous safety outcomes in acute care settings are only available for delegated exercise and mobility treatments for patients.^23,25^ A recent systematic review supports our findings, reporting no increased risk of harm to patients associated with a broad range of delegated AHA treatments occurring in hospital and community centres.^11^ These authors suggested healthcare organisations could be assured that AHAs can provide safe interventions under supervison.^9^ We were unable to collect more extensive safety data (e.g. intermittent desaturation with exercise) and this should be included in future research.

Generalisability of our findings should also be considered. Our robust methodology of using the Calderdale Framework to inform the inclusion of the AHA role was a deliberate strategy to optimise outcomes for the new AHA role and overall service delivery. Other studies developing AHA roles have not shown such successful outcomes^14,20^ and this may due to insufficient planning and training for all team members. Findings from this study suggest that delegated clinical and non-clinical roles could be established in other centres with similar education and training strategies.

It is possible that some changes observed in physiotherapist activity for inpatients may have been attributed to variations in the complexity of patients admitted to the CF centre across the two phases. There was no capacity to quantify patient complexity during each phase of the study. Variations in patient demand and complexity were minimised with data collection periods deliberately chosen to avoid peak holiday (December to January) and clinical demand (July to August). Additionally, data collection over a three-month period may not have been long enough to fully account for changes to service delivery and physiotherapists scope of practice. Further, there was some limitation to the activity codes used. Activity codes were selected in collaboration with the CF physiotherapy team which resulted in some limitation in our ability to tease out detailed information related to scope of practice.

Finally, it is important to acknowledge that most recently, provision of care to adults with CF is rapidly changing with the development of modulator therapies.^1^ These recent changes were unlikely to impact workloads for physiotherapists at this centre during the study period. Modulator therapies are demonstrating marked reductions in pulmonary exacerbations and consequent need for hospitalisation.^29,30^ The incidence of obesity and metabolic syndrome are increasing.^31,32^ This represents another role for delegated exercise testing and treatments. The COVID-19 pandemic has also affected models of care,^33^ with increased use of virtual platforms to deliver physiotherapy and other services. ^34^ Care is therefore being refocused to outpatient and virtual models.^33,34^ Establishing an AHA role that can perform safely delegated clinical tasks and potentially, many of the non-clinical tasks associated with virtual and face to face appointments, suggests even greater opportunity for this role to be used in current CF physiotherapy service provision.

This study describes the scope of practice undertaken by an AHA in an acute care service and the resultant changes to physiotherapy service provision within an adult CF centre. The AHA completed delegated clinical tasks such respiratory and exercise treatments and most of the exercise tests. AHA non-direct clinical care included managing equipment and patient related administration activities. Physiotherapist activity and scope of practice changed associated with provision of complex clinical care with increased activity in the clinics and undertaking more annual reviews. Physiotherapists also increased patient communication, management and research activity. Importantly, there were no safety issues reported. Critical to the successful establishment of the AHA role was the use of a workforce redesign tool to engage, develop, train and educate both the physiotherapists and the AHA for effective and safe delegation activities. There is potential for an AHA to enhance service delivery in other acute respiratory physiotherapy services.

## Data Availability

Data is part of current PhD process and further information can be requested through the corresponding author.

## Acknowledgements

The authors wish to sincerely thank the following people for their assistance with this study: Anthony Fish for his guidance and expertise in ABC data collection and management, Trent Donnelley for his expert physiotherapy care and to all other physiotherapists involved in the CF service during the study period

